# The changes of BMI in prediction of 10-year-hypertension risk in United States population- a historical cohort study

**DOI:** 10.1101/2024.01.22.24301636

**Authors:** Haoxuan Zou, Chumeng Wang, Wei Sun, Xiangqing Kong, Ming Jiang, Huayiyang Zou

## Abstract

**BACKGROUND:** This is a historical cohort study, utilizing data from the US National Health and Nutrition Examination Survey (NHANES) 2007-2018 to predict 10-year- hypertension-risk based on body mass index (BMI) variations.

**METHOD:** Participants aged 40-79 without a hypertension diagnosis 10 years before the baseline interview were included. Subjects were categorized based on five major BMI variation patterns. Various statistical analyses, including Chi-square test, T test, univariate and multivariate logistic regression, and P-trend analysis, were employed to assess hypertension incidence among groups. Restricted cubic splines (RCS) were used to examine the age-hypertension correlation.

**RESULTS:** Among 13,287 participants, Stable-Norm (maintaining normal BMI) exhibited the lowest 10-year hypertension risk. Other patterns—Max-OW (maximum BMI in overweight), OB-nOB (obese to non-obese), nOB-OB (non-obese to obese), and Stable-OB (maintaining obese)—showed increasing risks. Hypertension risk correlated quasi-linearly with age. Subgroups analysis suggested certain specific BMI variation modes and absolute weight change groups demonstrated equivalent risks to stable normal/weight groups, while others presented higher risks.

**CONCLUSION:** Maintaining normal BMI had the lowest 10-year hypertension risk, and returning to normal BMI showed equivalent risk. Weight gain remained a significant hypertension risk factor in US adults, particularly with advancing age.

## Introduction

Body mass index (BMI) serves as a key metric for evaluating weight status based on height, categorizing individuals into obesity (BMI ≥ 30 kg/m2), overweight (30 kg/m2 > BMI ≥ 25 kg/m2), normal (25 kg/m2 > BMI ≥ 19 kg/m2), and underweight (BMI < 19 kg/m2) [1]. As described in the guideline for the management of hypertension, numerous studies have identified obesity as an independent risk factor, contributing to 60% of essential hypertension cases [2]. Mechanisms underlying this association include overactive sympathetic nervous system, abnormal adipokine activation, and insulin resistance [2]. Previous trials have demonstrated significant reductions in systolic blood pressure (SBP) by 7–10 mm Hg and diastolic blood pressure (DBP) by 6–7 mm Hg following an approximate 8 kg weight loss. [3, 4]

Recent attention has shifted from BMI itself to BMI variations as a significant parameter[5]. Emerging research has linked BMI changes to cardiovascular disease (CVD) specific mortality[6]. However, the impact of BMI variations on hypertension prevalence remains underexplored. This study presents a historical cohort design aiming to assess the predictive value of diverse BMI variation patterns for the 10-year risk of hypertension in the adult U.S. population. Utilizing clinical data sourced from the National Health and Nutrition Examination Survey (NHANES), this research addresses an important gap in understanding the relationship between BMI variations and hypertension.

## Method

### Study population

Our study population was derived from NHANES spanning from 2007 to 2018. The baseline was determined as the interview time for questionnaires. The inclusion criteria was defined as 1) age of 40-79 at baseline. 2) individuals without a history of hypertension within the 10 years preceding the baseline assessment. Exclusion criteria comprised the subjects with crucial missing data (eg. weight, height, hypertension diagnosis).

### Assessments of BMI variation and covariates

Weight information at age 25 and 10 years prior to baseline was obtained through the NHANES questionnaire, while baseline height and weight were measured during mobile physical examinations. BMI at age 25 and 10 years before baseline was computed as weight (kg) divided by the square of height (m2). BMI categories were defined as underweight (≤18.4), normal weight (18.5-24.9), overweight (25.0-29.9), and obese (≥30.0) [7] Covariate data, encompassing age, gender, race, educational level, marital status, smoking history, alcohol use, sodium intake, physical activity status, and self-reported incidences of high blood cholesterol and diabetes, were sourced from NHANES baseline questionnaires and examinations.

BMI variations between the two time points were categorized into five major patterns: stable normal pattern (Stable-Norm, BMI<25.0 at both times), maximum overweight pattern (Max-OW, BMI 25.0-29.9 at either time but not ≥30.0 at the other time), obese to non-obese pattern (OB-nOB, BMI≥30.0 at younger age and <30.0 later), non-obese to obese pattern (nOB-OB, BMI<30.0 at younger age and ≥30.0 later), and stable obesity pattern (Stable-OB, BMI≥30.0 at both times). The method has already been applied in previous studies [8, 9]. We also demonstrated the correlations of 16 specific BMI change modes (BMI change among underweight, normal, overweight, obese) and absolute weight change modes with the incidence of hypertension respectively. Absolute weight change modes were defined as weight loss (weight loss >2.5 kg), the stable weight (weight change within 2.5 kg), the small to moderate weight gain (weight gain ≥2.5 kg and <10 kg), the moderate to large weight gain (weight gain ≥10 kg and <20 kg), and the extreme weight gain (weight gain ≥20 kg) [10].

### Ascertainment of hypertension

The prevalence of hypertension was ascertained using data from the Blood Pressure & Cholesterol Questionnaire of NHANES 2007-2018. Given the potential influence of drug control on hypertension, our study outcome depended on participants’ responses to question BPQ020, specifically, whether they had been informed by a doctor or health professional of a hypertension diagnosis or prescribed anti-hypertensive drugs. This methodology aligns with a previously conducted study, ensuring consistency and comparability in our approach[11].

### Statistical analysis

Variables adhering to normal distribution were expressed as means and standard deviations, while those deviating from normal distribution were presented as medians and interquartile ranges. Categorical variables were represented as counts and percentages.

Initially, differences in baseline characteristics among the five BMI variation patterns were assessed. For categorical variables, the χ2 test was employed, and for continuous variables conforming to normal distribution, multivariate variance analysis was utilized; otherwise, the Wilcoxon rank sum test was applied. The correlation between BMI at the two time points and the incidence of hypertension was examined using Spearman’s correlation test respectively, followed by P trend analysis.

Logistic regression was employed to calculate the 10-year odds ratios (OR) and their corresponding 95% confidence intervals (CI) for hypertension incidence. Additionally, non-parametric restricted cubic splines (RCS) were used to explore the potential non-linear correlation between age and hypertension prevalence. In subgroup analysis, the same approaches were applied to investigate the correlation between specific BMI variation and absolute weight change modes with hypertension prevalence.

For the analysis of the five major BMI variation patterns, a multivariate logistic model was employed to explore the association between BMI variation and the prevalence of hypertension in four sequential models. Model 1 involved no adjustments. In Model 2, adjustments were made for baseline age (year), sex, race (non-Hispanic white, non-Hispanic black, Mexican American, and others), education (Less than high school, High school or equivalent, College or above), marital status (Married, Widowed, Divorced, Separated, Never married, Living with partner). Model 3 further included adjustments for the incidence of diabetes (not diagnosed with diabetes, diagnosed with diabetes, on the borderline of diabetes) and hypercholesterolemia (not diagnosed with hypercholesterolemia, diagnosed with hypercholesterolemia). Model 4 extended adjustments to incorporate smoking status (ever smoked 100 cigarettes or more), drinking status (average drinks per day: non-drinker, low to moderate drinker defined as drinking <1 drink/day in women and <2 drinks/day in men, heavy drinker defined as ≥1 drink/day in women and ≥2 drinks/day in men) [12], sodium intake status(high sodium intake defined by the World Health Organization as >5 g sodium per day), and insomnia condition (diagnosed with insomnia or not). Additionally, baseline physical activities (engagement in vigorous/moderate recreational activities) during leisure time was considered potential confounders. The data were analyzed using Recovery Component version 4.3.1.

### Patient and public involvement

No subjects were engaged in the design, execution, questionnaire settings, or outcome measurements of our study.

## Result

### Baseline characteristics and BMI variation groups

A total of 60,634 participants were initially screened from NHANES (2007-2018), and ultimately, 13,287 participants were enrolled. Among them, 4,232 subjects developed hypertension in the past 10 years from baseline. In the enrolled participants, males (49.4%) and females (50.6%) were represented in equivalent proportions. Among the five major BMI variation patterns, 38.5% of participants belonged to the Stable-Norm group, 36.2% to the Max-OW group, 0.9% to the OB-nOB group, 18.6% to the nOB-OB group, and 5.8% to the Stable-OB group. Further details are presented in Table 1.

**Table 1.**
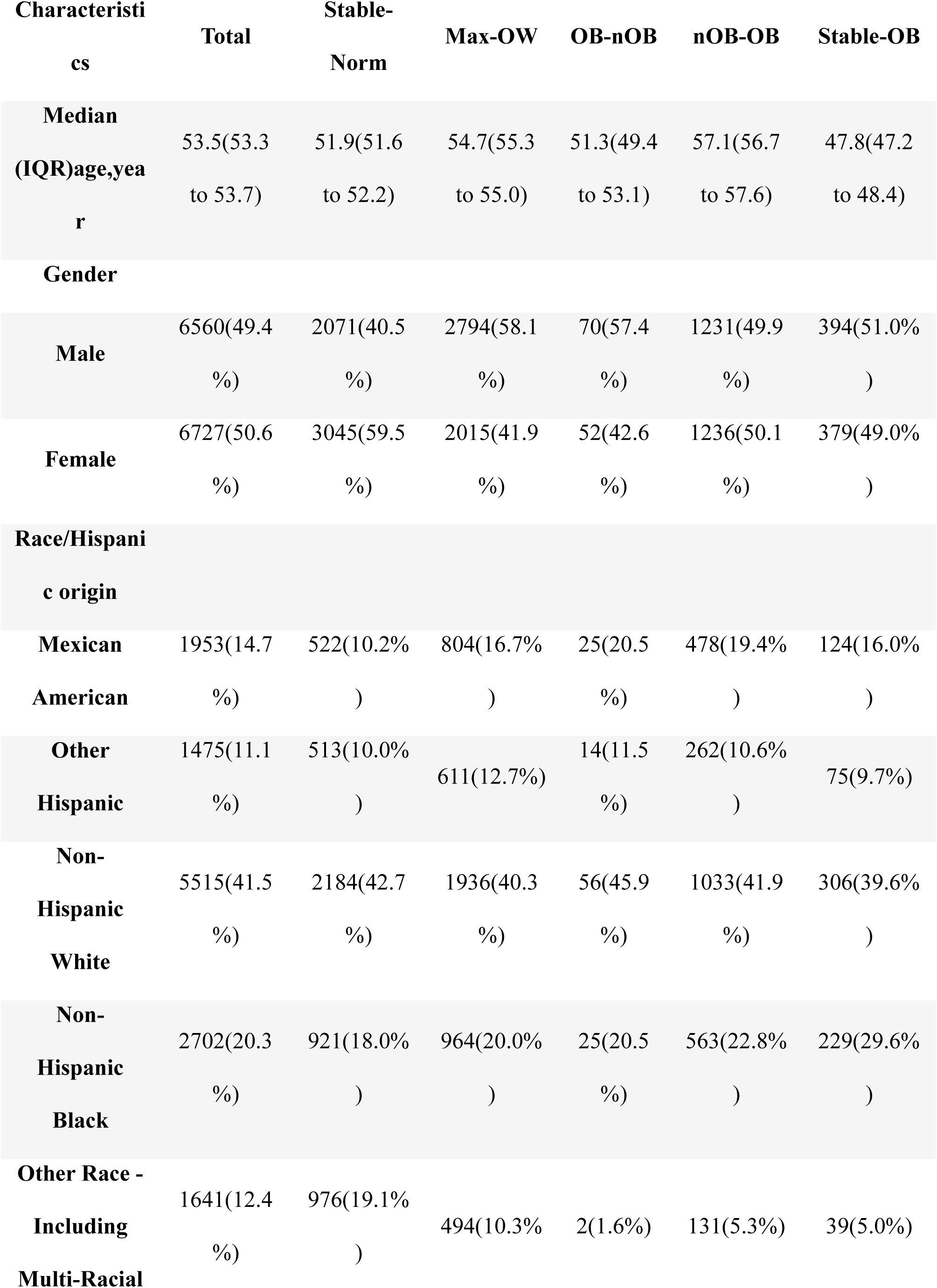
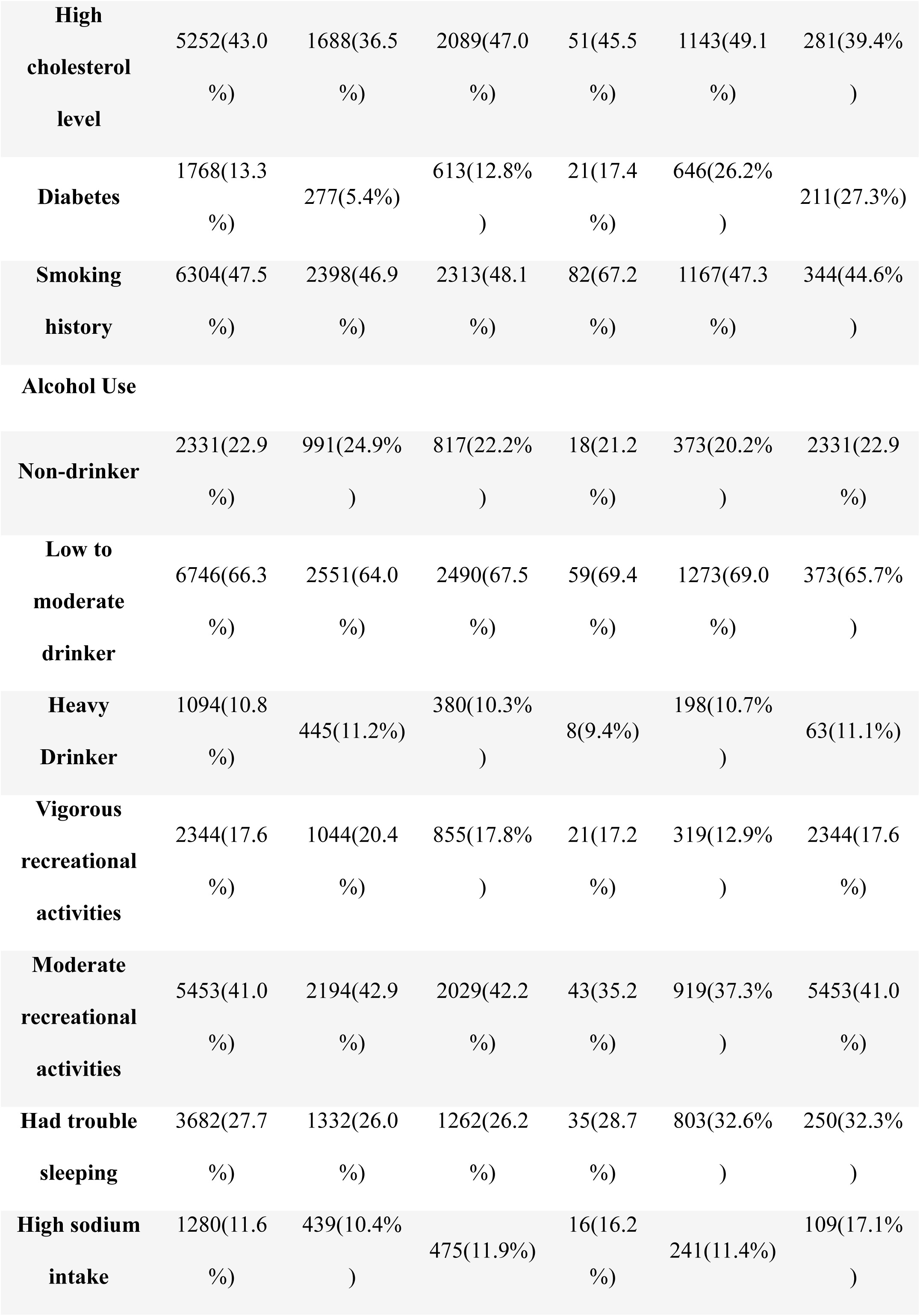
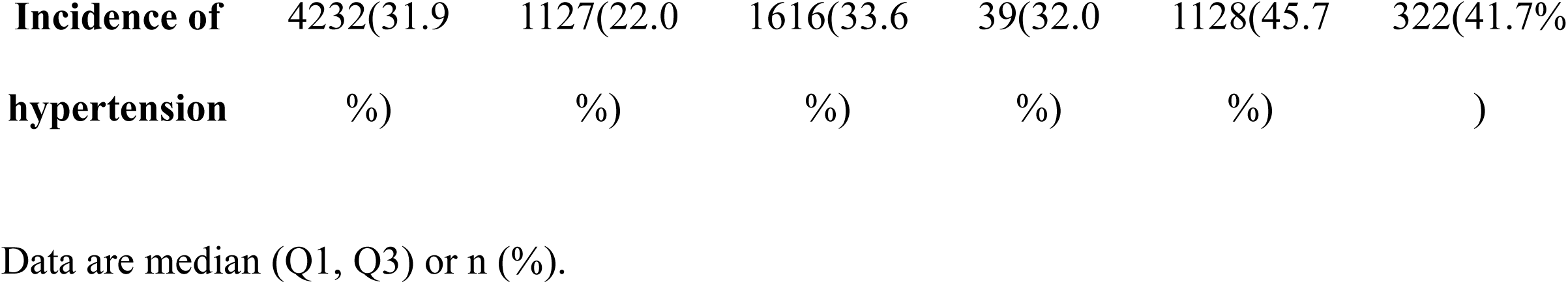
Basline characteristics of study participants aged 40-79 in NHANSE 2007-2018 according to BMI variation patterns from age 25 to 10 years before baseline.

### Correlations of major BMI variation patterns with incidence of hypertension

Spearman’s correlation test results revealed a positive correlation between BMI at two time points (age 25 and 10 years before baseline) and the incidence of hypertension (Age 25: r=0.064, P<0.001; 10 years before baseline: r=0.193, P<0.001). These findings align with the results of P trend analysis, indicating a significant positive trend for both (Age 25: P<0.001; 10 years before baseline: P<0.001) [Table 2].

**Table 2.** Correlations between BMI at age 25/10 years before baseline and prevalence of hypertension.

| Covariates | Spearman's correlation test |  | P trend analysis |
| --- | --- | --- | --- |
|  | Correlation coefficient | P value | P value |
| BMI at age 25 | 0.064 | <0.001 | <0.001 |
| BMI 10 years before<br>baseline | 0.193 | <0.001 | <0.001 |

In Model 4, encompassing all participants and utilizing the Stable-Norm group as the reference, the Max-OW group exhibited a higher incidence rate of hypertension (OR 1.58, 95% CI 1.44-1.74, P<0.001). Similarly, the nOB-OB group (OR 2.25, 95% CI 2.01-2.52, P<0.001), Stable-OB group (OR 2.32, 95% CI 1.96-2.76, P<0.001), and OB-nOB group (OR 1.49, 95% CI 0.99-to 2.23, P=0.057) were associated with elevated hypertension incidence [Table 3 and Figure 1]. Consistent trends were observed in Model 1-3 [Figure 2].

**Figure 1.**
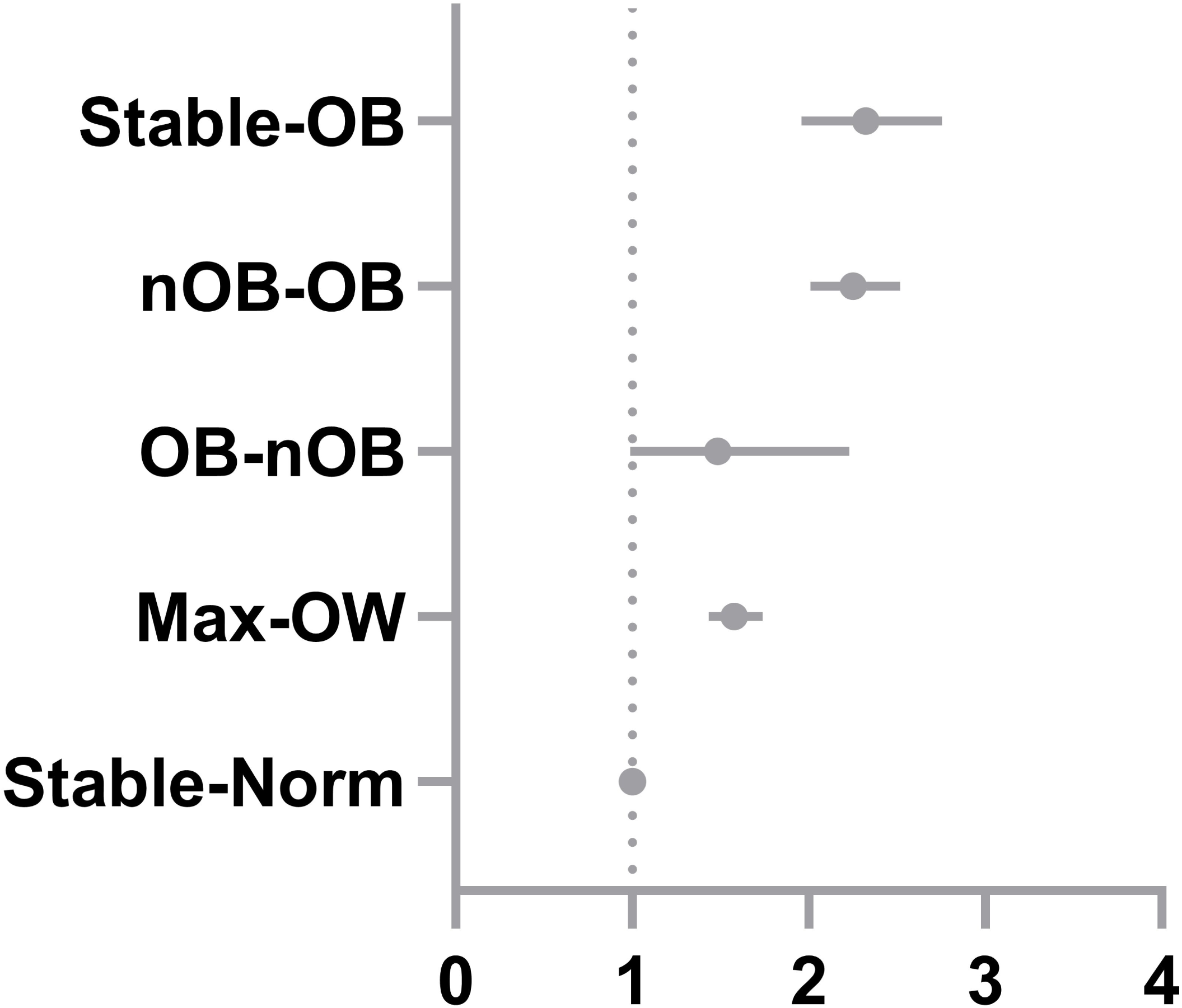
Forest plots of odds ratios regarding the risk of hypertension of BMI variation patterns in NHANSE 2007-2018. The x-coordinate of this figure was defined as odds ratios. Solid points represent estimates of odds ratios and solid lines represent 95% CIs, both were obtained using logistic regression. Stable-Norm, stable normal pattern (BMI<25.0 at both times); Max-OW, maximum overweight pattern (BMI 25.0-29.9 at either time but not ≥30.0 at the other time); OB-nOB, obese to non-obese pattern (BMI≥30.0 at younger age and <30.0 later); Stable-OB, stable obesity pattern (BMI≥30.0 at both times).

**Figure 2.**
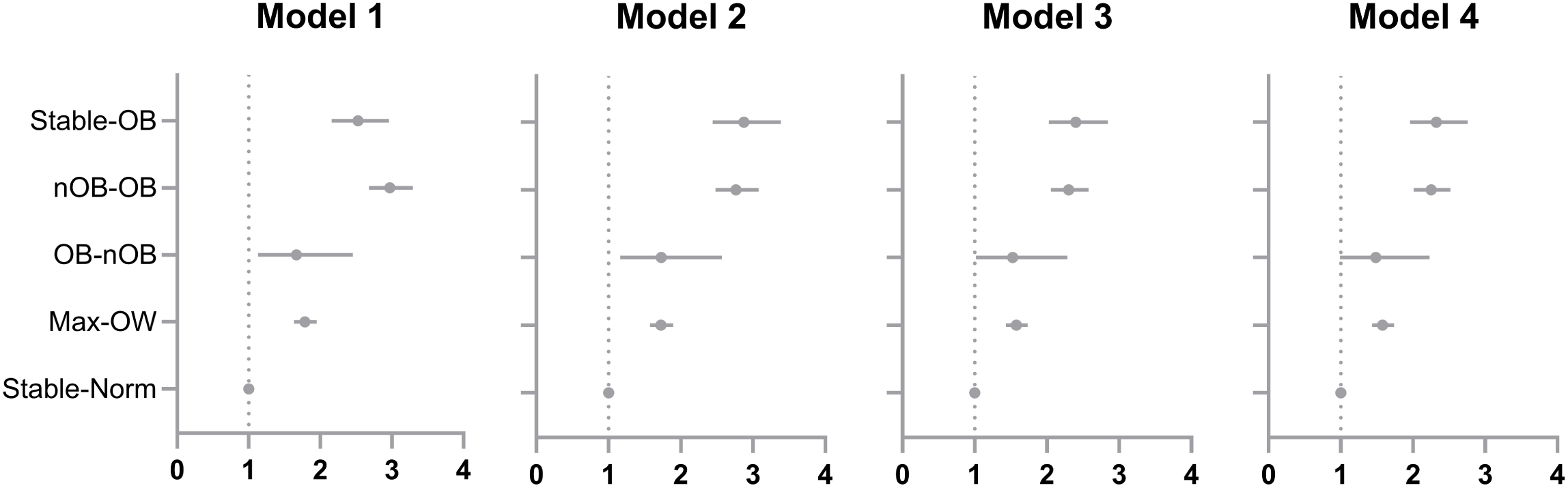
Forest plots of odds ratios regarding the risk of hypertension of BMI variation patterns in model 1-4. The x-coordinate of this figure was defined as odds ratios. Solid points represent estimates of odds ratios and solid lines represent 95% CIs, both were obtained using logistic regression. Stable-Norm, stable normal pattern (BMI<25.0 at both times); Max-OW, maximum overweight pattern (BMI 25.0-29.9 at either time but not ≥30.0 at the other time); OB-nOB, obese to non-obese pattern (BMI≥30.0 at younger age and <30.0 later); Stable-OB, stable obesity pattern (BMI≥30.0 at both times).

**Table 3.** Odds ratios of hypertension with BMI change patterns.

|  | Groups | Sample size | Odds ratios | P values |
| --- | --- | --- | --- | --- |
| **Major BMI change patterns** | Stable-Norm | 5112 | 1 |  |
|  | Max-OW | 4797 | 1.58 (1.44 to 1.74) | P<0.001 |
|  | OB-nOB | 122 | 1.49 (0.99 to 2.23) | P=0.057 |
|  | nOB-OB | 2459 | 2.25 (2.01 to 2.52) | P<0.001 |
|  | Stable-OB | 772 | 2.32 (1.96 to 2.76) | P<0.001 |
| **Specific BMI change modes** | Stable normal | 4293 | 1 |  |
|  | Normal to overweight | 3020 | 1.69 (1.51 to 1.88) | P<0.001 |
|  | Normal to obese | 1124 | 2.18(1.87 to 2.53) | P<0.001 |
|  | Overweight to normal | 161 | 0.88 (0.58 to 1.33) | P=0.542 |
|  | Stable overweight | 1444 | 1.58 (1.36 to 1.82) | P<0.001 |
|  | Overweight to obese | 1272 | 2.46 (2.13 to 2.84) | P<0.001 |
|  | Obese to normal | 31 | 0.92(0.39 to 2.21) | P=0.854 |
|  | Obese to overweight | 91 | 1.79 (1.13 to 2.83) | P=0.014 |
|  | Stable obese | 772 | 2.39 (2.01 to 2.84) | P<0.001 |
|  | Stable underweight | 184 | 0.84 (0.56 to 1.26) | P=0.391 |
|  | Underweight to normal | 576 | 1.32(1.07 to 1.61) | P=0.008 |
|  | Underweight to overweight | 167 | 1.84 (1.32 to 2.55) | P<0.001 |
|  | Underweight to obese | 63 | 2.51 (1.48 to 4.25) | P=0.001 |
|  | Normal to underweight | 59 | 1.17 (0.63 to 2.17) | P=0.631 |
|  | Overweight to underweight | 5 | insufficient sample size |  |
| **Weight change patterns** | Weight change within 2.5kg | 2967 | 1 | - |
|  | Weight loss >2.5kg | 818 | 0.99 (0.82 to 1.20) | P=0.95 |
|  | 2.5kg≤Weight gain <10kg | 4498 | 1.26 (1.12 to 1.41) | P<0.001 |
|  | 10kg≤Weight gain<20kg | 3099 | 1.69 (1.50 to 1.91) | P<0.001 |
|  | 20kg≤Weight gain | 1880 | 2.11 (1.85 to 2.42) | P<0.001 |

Stable-Norm, stable normal pattern (BMI<25.0 at both times); Max-OW, maximum overweight pattern (BMI 25.0-29.9 at either time but not $\geq 30.0$ at the other time); OB-nOB, obese to non-obese pattern (BMI $\geq 30.0$ at younger age and <30.0 later); Stable-OB, stable obesity pattern (BMI $\geq 30.0$ at both times). P values of different BMI/weight change patterns were obtained using logistic regression.

Further exploration of the RCSs between age and the prevalence of hypertension across the five major BMI variation patterns revealed a generally positive quasi-linear trend in the overall status, with the medium OR observed at age 55. Similar trends were identified in the Stable-Norm and nOB-OB groups, showcasing ORs of 1 at ages 53 and 58, respectively. In contrast, the Max-OW group exhibited an inverted U-shaped parabolic trend, reaching its peak hypertension risk at age 67. Both the OB-nOB and Stable-OB groups displayed multi-segment trends, with the peak point at age 55 for Stable-OB and a valley point at age 43. The OB-nOB group demonstrated a W-shaped trend, featuring a higher turning point at age 53 and two valley points at ages 47 and 64 [Figure 3].

**Figure 3.**
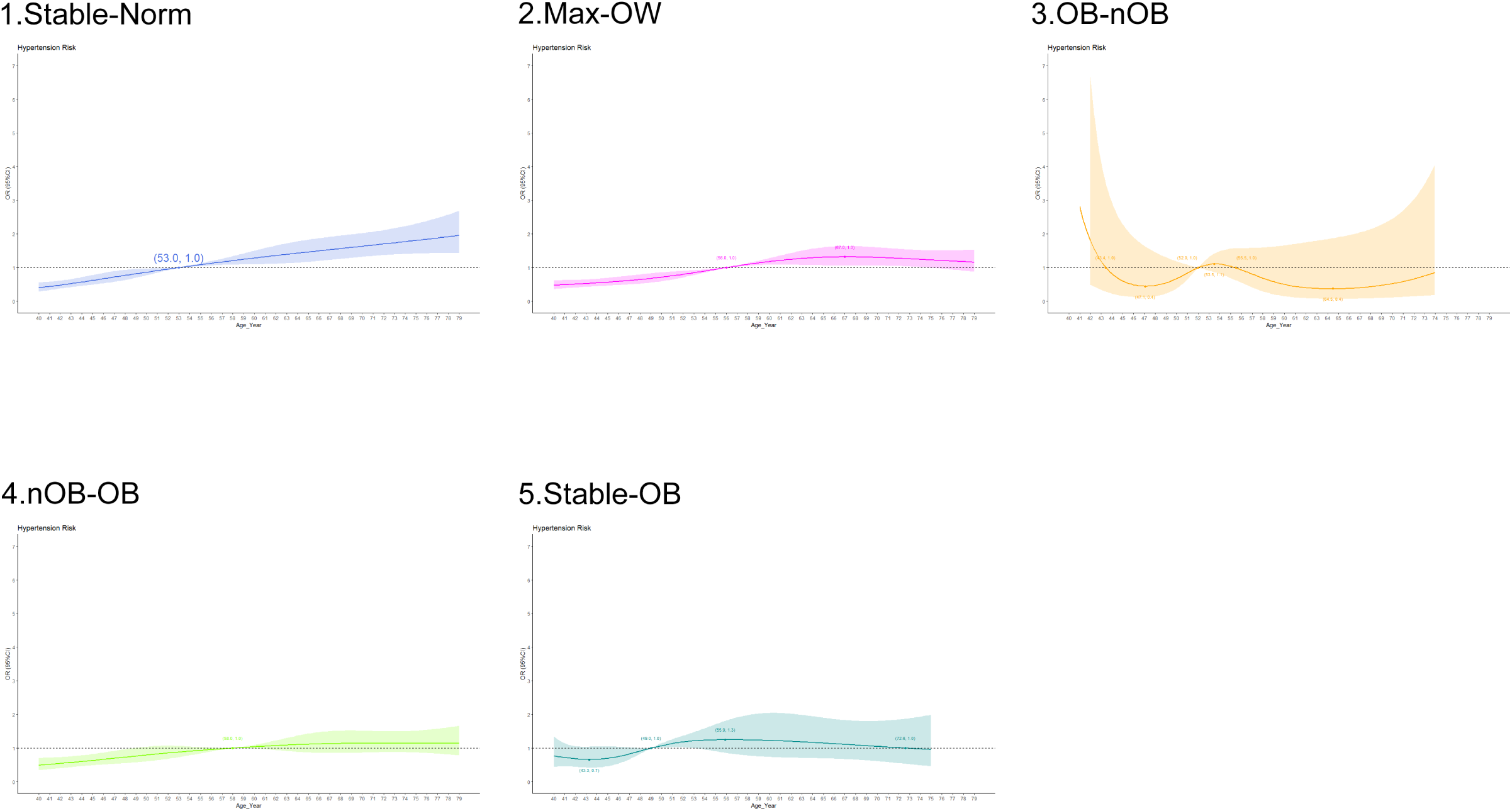
the RCS between age and the risk of hypertension in BMI variation patterns respectively. The x-coordinate of each figure was defined as baseline age (years) and the y-coordinate was defined as odds ratios. Solid lines represent estimates of odds ratios and colored parts represent 95% CIs. Stable-Norm, stable normal pattern (BMI<25.0 at both times); Max-OW, maximum overweight pattern (BMI 25.0-29.9 at either time but not ≥30.0 at the other time); OB-nOB, obese to non-obese pattern (BMI≥30.0 at younger age and <30.0 later); Stable-OB, stable obesity pattern (BMI≥30.0 at both times).

### Correlations of specific BMI variation modes and absolute weight change with incidence of hypertension

Specific BMI change modes, representing weight status changes between two time points (usually age 25 to 10 years before baseline), were investigated, excluding the obese to underweight group due to a lack of eligible samples. Utilizing the stable normal (normal to normal) group as a reference, higher prevalence of hypertension was observed in the underweight to normal (OR 1.32, 95% CI 1.07-1.61, P=0.008), underweight to overweight (OR 1.84, 95% CI 1.32-2.55, P<0.001), underweight to obese (OR 2.51, 95% CI 1.48-4.25, P=0.001), normal to overweight (OR 1.69, 95% CI 1.51-1.88, P<0.001), normal to obese (OR 2.18, 95% CI 1.87-2.53, P<0.001), stable overweight (OR 1.58, 95% CI 1.36-1.82, P<0.001), overweight to obese (OR 2.46, 95% CI 2.13-2.84, P<0.001), obese to overweight (OR 1.79, 95% CI 1.13-2.83, P=0.014) and stable obese (OR 2.39, 95% CI 2.01-2.84, P<0.001) groups, whereas the stable underweight (OR 0.84, 95% CI 0.56-1.26, P=0.391), normal to underweight (OR 1.17, 95% CI 0.63-2.17, P=0.631), overweight to normal (OR 0.88, 95% CI 0.58-1.33, P=0.542) and obese to normal (OR 0.92, 95% CI 0.39-2.21, P=0.854) groups showed no difference. [Table 3 and Figure 4].

**Figure 4.**
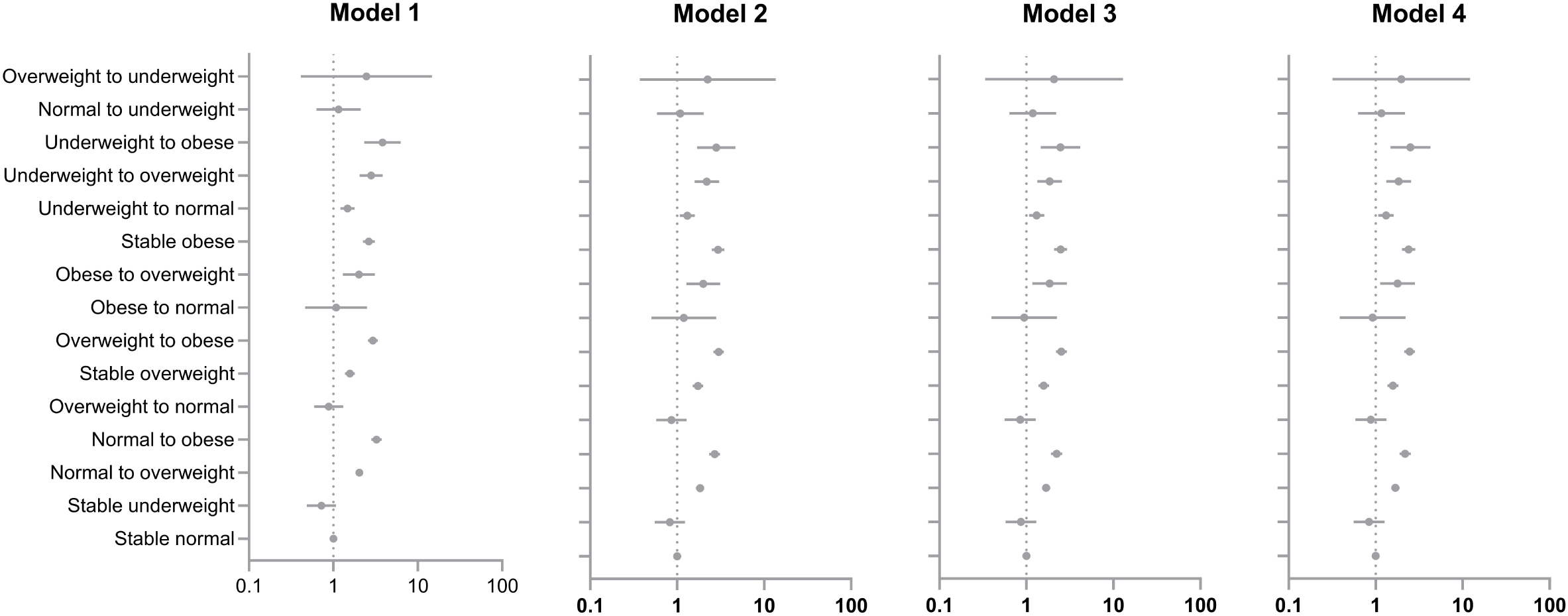
Forest plots of odds ratios regarding the risk of hypertension of specific BMI variation patterns in model 1-4. The x-coordinate of this figure was defined as odds ratios. Solid points represent estimates of odds ratios and solid lines represent 95% CIs, both were obtained using logistic regression.

Regarding absolute weight changes, in comparison to the stable weight group, the weight loss group exhibited no significant difference in the risk of hypertension (OR 0.99, 95% CI 0.82-1.20, P=0.95). However, the small to moderate weight gain group (OR 1.26, 95% CI 1.12-1.41, P<0.001), the moderate to large weight gain group (OR 1.69, 95% CI 1.50-1.91, P<0.001), and the extreme weight gain group (OR 2.11, 95% CI 1.85-2.42, P<0.001) all demonstrated higher risks of hypertension, suggesting a positive correlation between absolute weight gain and the risk of hypertension [Table 3 and Figure 5].

**Figure 5.**
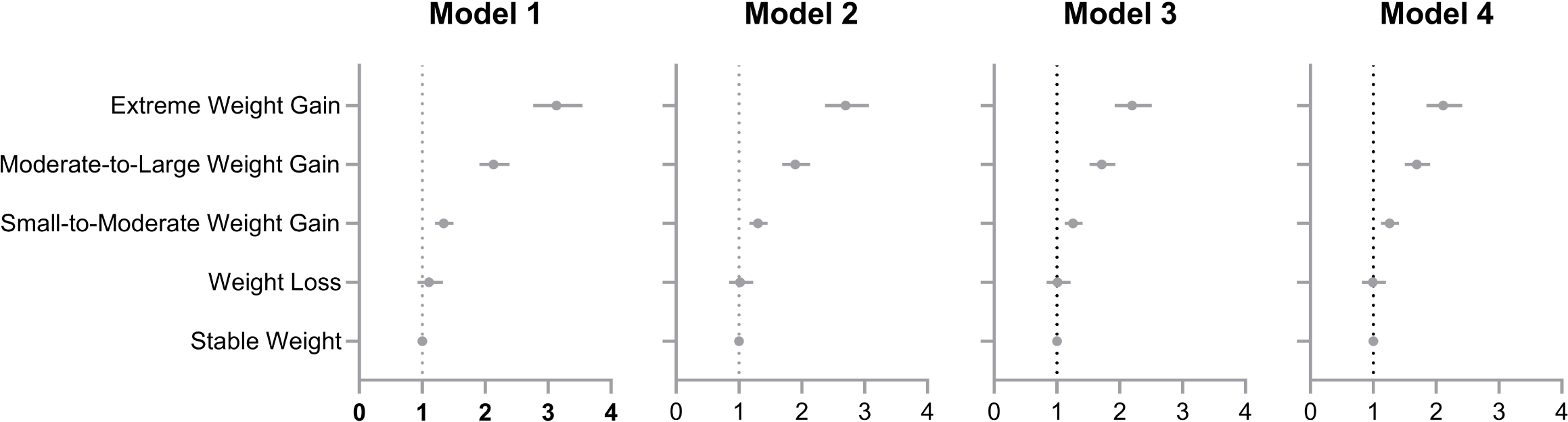
Forest plots of odds ratios regarding the risk of hypertension of absolute weight change patterns in model 1-4. The x-coordinate of this figure was defined as odds ratios. Solid points represent estimates of odds ratios and solid lines represent 95% CIs, both were obtained using logistic regression.

## Discussion

This study introduced various BMI change modes to predict the 10-year hypertension risk, revealing distinct associations between different patterns and hypertension incidence. The findings indicated that maintaining a normal BMI status was linked to the lowest hypertension risk among major BMI variation patterns, followed by OB-nOB, Max-OW, nOB-OB, and Stable-OB. In specific BMI change modes, keeping normal BMI still possessed lowest hypertension risk, however, transitions such as overweight to normal, obese to normal exhibited equivalent risks with the stable normal group and transitioning from obese to overweight still presented a higher risk (albeit lower than stable obese), which suggested that weight loss may offer protective effects, and the maximum protective effects achieved when BMI returns to normal. Furthermore, decreasing BMI from normal to underweight did not generate additional benefits. Regarding absolute weight change modes, weight loss exceeding 2.5kg demonstrated equivalent risk with the stable weight group, while the others showed elevated risks. These findings emphasizing the importance of both initial BMI status and subsequent changes for long-term health outcomes.

The whole trend of hypertension risk with age in our study was positively quasi-linear correlated as the various studies suggested [13, 14]. The Stable-Norm, nOB-OB, Max-OW, and Stable-OB groups also exhibited positively trending patterns with age.

However, the OB-nOB group presented a unique negatively multi-segmental trend with W-shaped fluctuation in the curve. Proper reasons might involve that diverse weight loss methods may yield varying effects, and the corresponding impact and duration of blood pressure reduction may not be consistent [15]. Additionally, relatively small sample size, with only 122 subjects in the OB-nOB group could also disturb the result.

Many studies have delved into the association between BMI and the prevalence of hypertension [7, 11, 15, 16], pointing out that BMI is non-negligible in the prediction of hypertension incidence. This study extends the exploration by investigating the association between BMI variation and the 10-year risk of hypertension. The meticulous classification of BMI variation, along with comprehensive adjustments for potential confounders related to lifestyle habits and basic disease history enhances the validity of the study results. While prior research predominantly focused on the elderly population [11, 17–19], our study inclusively considers adults across a broader age range, examining the impact of early BMI on subsequent hypertension incidence. This approach expands the understanding of the complex interplay between BMI variations and long-term hypertension risk in a diverse adult population.

Among the hypertension confounders considered in this study, there was no significant difference in hypertension risk between males and females. This study revealed a significantly higher risk of hypertension among non-Hispanic Black participants, while the remaining racial groups showed no difference compared to Mexican Americans. Traditional hypertension risk factors such as hypercholesterolemia, low physical activity, and diabetes (including borderline diabetes) revealed significantly higher risks, consistent with established literature [7]. However, smoking was associated with a milder increase in risk compared to previous reports [20, 21]. It is noteworthy that, contrary to traditional risk factors [18, 22, 23], high sodium intake and drinking showed no difference in increasing hypertension risk in this study [Table 4]. A potential explanation could be the relatively short diet history (12 months) recorded before the interview. Besides traditional risk factors, insomnia emerged as a novel risk factor, contributing to an additional 71% hypertension risk compared to non-insomnia individuals in this study. The possible mechanisms underlying this association may be linked to the over-activated sympathetic nervous system, as suggested in recent studies [24, 25]. These findings emphasize the multifaceted nature of hypertension risk factors and highlight the importance of considering emerging factors in comprehensive risk assessments.

**Table 4.**
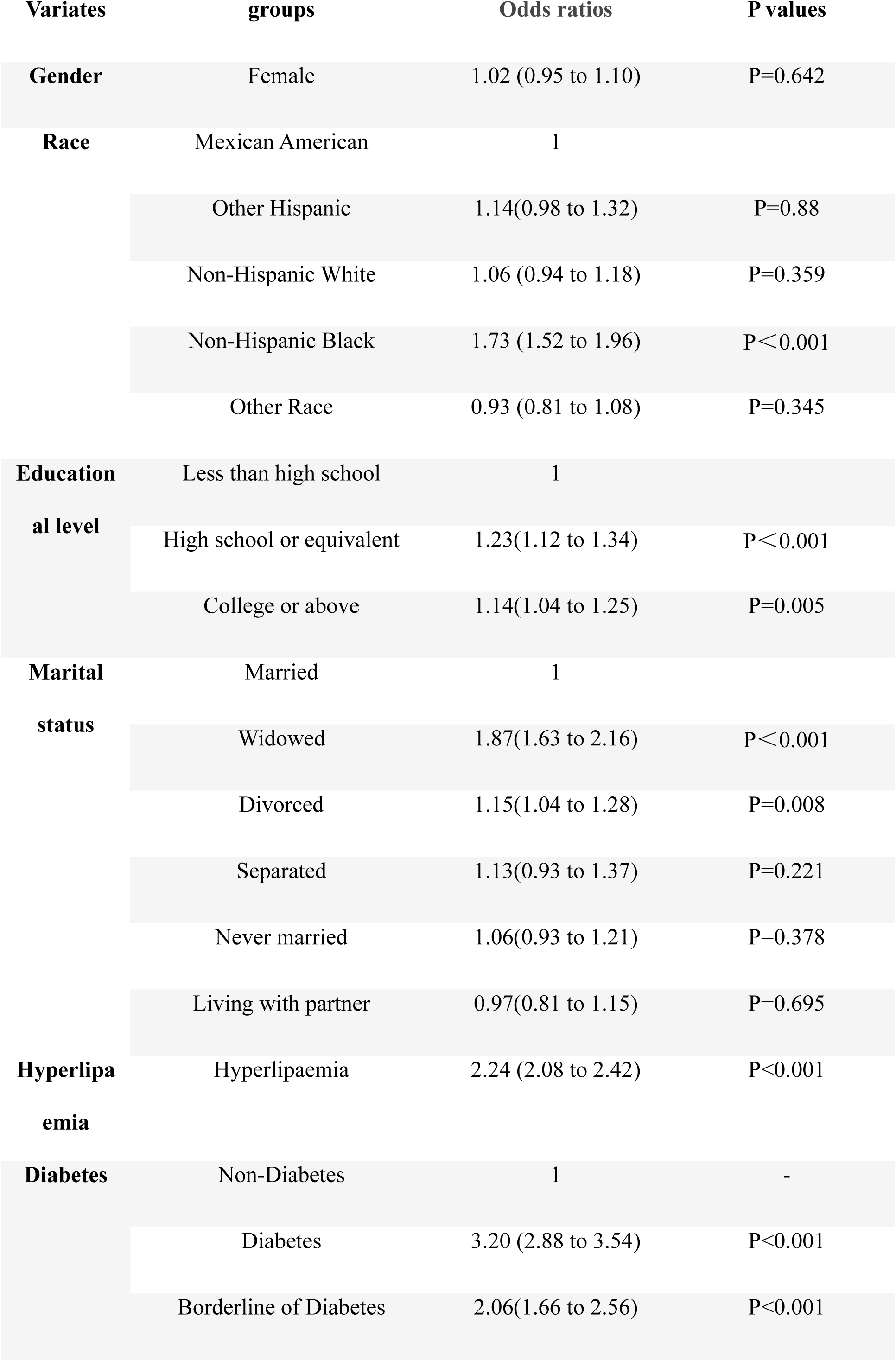
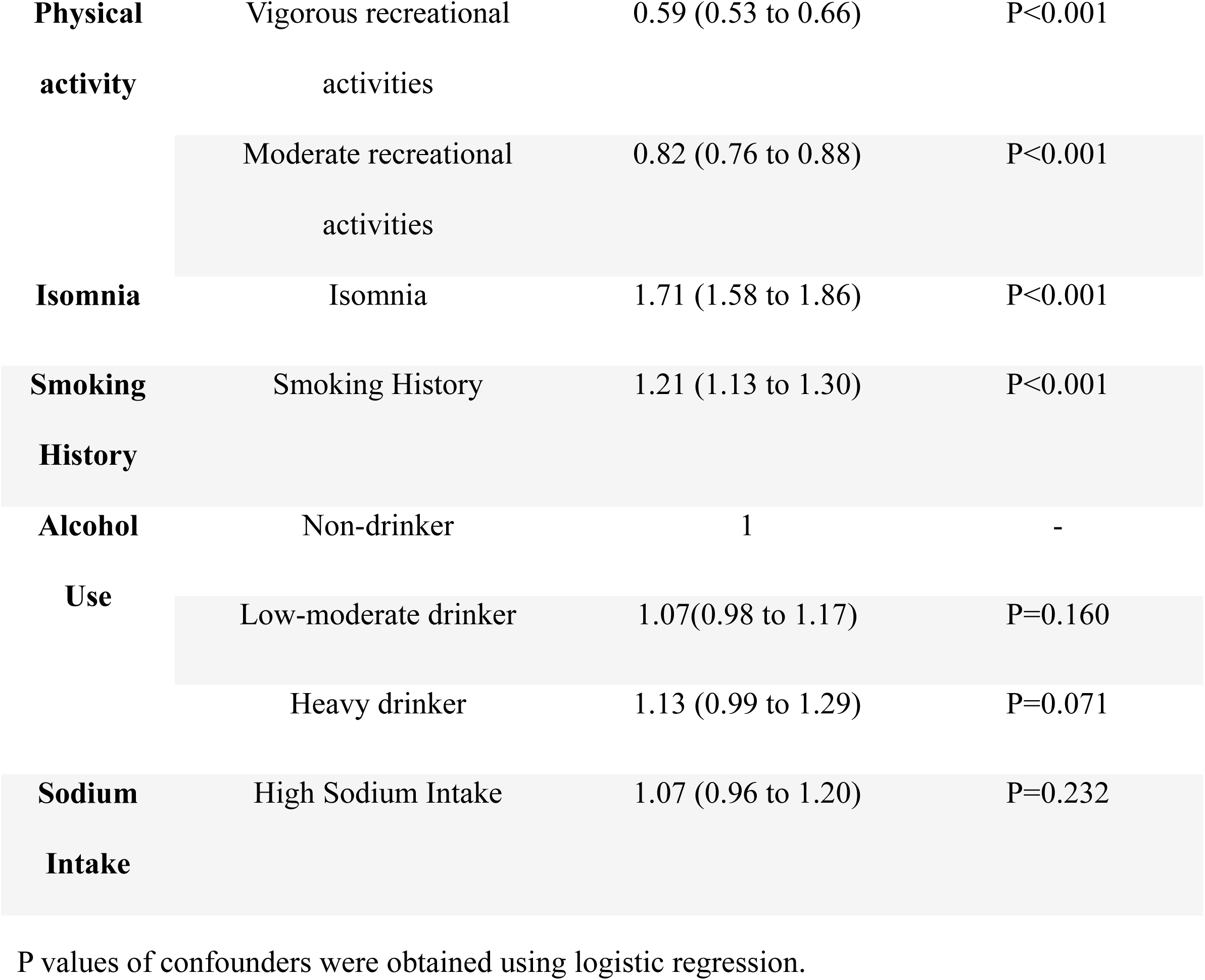
Odds ratios of hypertension with confounders.

## Limitations

This study had several limitations. Firstly, the ascertainment of our study relies on self-reported medical history, and the weight at age 25 and 10 years prior to baseline was recalled by participants rather than accurately measured in the past, which might introduce misclassification bias. Secondly, we could not distinguish intentional BMI loss from BMI loss caused by disease or aging and we assume that the height was stable during the adulthood which might be influenced by hunchback and osteoporosis, particularly in the elderly. Thirdly, our study had excluded the participants who didn’t meet the inclusion criteria, making the large sample of our study no longer demographically representative. Finally, some specific BMI variation pattern groups have small sample sizes, potentially impacting the accuracy of data analysis. Future NHANSE surveys incorporating more precise indexes associated with body size (eg. waist circumstances) would strengthen the validity and specificity of researches on the predictive value of BMI variations in the prevalence of hypertension.

## Conclusion

The study suggests that maintaining a normal BMI is associated with the lowest 10-year hypertension risk. Reverting BMI back to normal may confer an equivalent 10-year hypertension risk compared to consistently maintaining a normal BMI. Weight gain remains a significant risk factor for hypertension in US adults, and this influence appears to amplify with age. Further research incorporating more specific and comprehensive data on potential confounders is needed to validate and expand upon these findings.

## Data Availability

Data of our article originated from the US National Health and Nutrition Examination Survey (NHANES).

https://wwwn.cdc.gov/nchs/nhanes/Default.aspx

## Acknowledgments

none

